# Incidence and Predictors of IOP-Lowering Treatment Following Detection of Referable Glaucoma in a Teleretinal Screening Program

**DOI:** 10.64898/2026.06.02.26354782

**Authors:** Kyle Bolo, Brandon J. Wong, Jiun L. Do, Jose-Luis Ambite, Zhiwei Li, Carl Kesselman, Lauren P. Daskivich, Benjamin Y. Xu

## Abstract

**Purpose:** To evaluate the incidence and baseline predictors of intraocular pressure (IOP)-lowering treatment following detection of referable glaucoma by teleretinal screening.

**Design:** Retrospective cohort study.

**Methods:** Participants were derived from a safety-net teleretinal diabetic retinopathy screening program (2013-2024). Participants included individuals who screened positive for referable glaucoma (cup-to-disc ratio [CDR] ≥0.6 or CDR asymmetry ≥0.2) and completed in-office diagnostic evaluation. The primary outcome was initiation of IOP-lowering treatment (medication, laser, or surgery) and the secondary outcome was intervention with surgery. Cumulative incidence functions were estimated, accounting for loss to follow-up. Fine-Gray models were used to identify baseline screening predictors to risk stratify each outcome. Glaucoma diagnosis was approximated using diagnostic codes and chart review.

**Results:** 2,367 participants were included. The cumulative incidence of treatment was 19.6% (95% CI: 18.0-21.2) at Year 1 and 45.1% (42.1-48.1) at Year 8. Early treatment occurred primarily in glaucoma cases, whereas treatment accumulated longitudinally in glaucoma suspects, reaching 36.5% (31.6-41.5) by Year 8. Surgery was less common (8-year incidence: 5.3%). Baseline screening data predicted treatment and surgery, enabling risk stratification. At Year 8, cumulative incidence differed substantially between high- and low-risk groups (treatment: 59.9% vs. 31.2%; surgery: 9.7% vs. 1.0%). Older age (sub-distribution hazard ratio [SHR] 1.03 per year, p<0.001), Black race (SHR 1.50, p<0.001), and personal history of glaucoma (SHR 1.90, p<0.001) were associated with treatment; Asian race was protective (0.71, p=0.03). Older age (SHR 1.06, p<0.001), worse visual acuity (SHR 5.11 per logMAR unit, p<0.001), and screening at a hospital-based site (SHR 2.46, p=0.003) were associated with surgical treatment.

**Conclusion:** Nearly half of safety-net diabetic patients screening positive for referable glaucoma initiated IOP-lowering treatment over 8 years, while few received surgery. Baseline screening characteristics enabled risk stratification of treatment and surgery. These findings address an evidence gap about longitudinal consequences of screening and suggest that its impact extends beyond detection of prevalent glaucoma to include identification of high-risk glaucoma suspects who warrant ongoing surveillance.

**Highlights:**

- Longitudinal treatment outcomes address consequences of glaucoma screening
- Nearly half of diabetic patients with referable glaucoma received treatment
- Few patients progressed to surgery
- Characteristics assessed at screening enabled risk stratification

## Introduction

Glaucoma is a leading cause of irreversible blindness worldwide.^1^ More than half of glaucoma cases remain undiagnosed globally, including an estimated 61.9% in the United States (U.S.).^2^ The current paradigm of glaucoma detection in the U.S. relies on case finding through comprehensive eye examinations, which may be prompted by visual symptoms or referral from a primary care clinician or optometrist for evaluation of risk factors.^3^ This approach often fails to identify early, asymptomatic disease, when intervention may be most effective. Population-based screening may improve identification of glaucoma prior to severe visual morbidity and has been implemented by several large U.S. healthcare systems, including the Veterans Health Administration.^4,5^ Therefore, evaluating downstream clinical outcomes of screened populations is critical to determine the true impact of screening programs and inform policy recommendations; however, data on the long-term clinical consequences of glaucoma screening remain limited, highlighting a need for longitudinal evidence in this setting.

Multiple population-based studies have evaluated glaucoma screening.^6–10^ The ongoing, prospective SIGHT trials have preliminarily demonstrated that targeted screening protocols can improve program yield and will assess interventions to increase engagement with follow-up.^11,12^ Despite these advances, the U.S. Preventive Services Task Force (USPSTF) has concluded there is insufficient evidence to support population-based glaucoma screening.^13^ One evidence gap is the limited data on longitudinal clinical outcomes among patients after screening. Most trials focus on diagnostic endpoints at the initial post-screening evaluation and do not capture long-term clinical endpoints, such as the initiation of intraocular pressure (IOP)-lowering treatment, changes in IOP, or progression of structural and functional disease—outcomes identified in USPSTF key questions.^14,15^ Given the high proportion of glaucoma suspects identified in screening programs (up to 29.2%), reliance on proximal diagnostic endpoints may underestimate the long-term benefit of screening, particularly for identifying high-risk individuals who subsequently require treatment.^14^

In this study, we investigated the longitudinal incidence of IOP-lowering treatment after screening positive for referable glaucoma in the Los Angeles County Department of Health Services (LAC DHS) teleretinal screening program.^16^ As the second largest municipal healthcare system in the U.S., LAC DHS screens approximately 1,750 patients per month through a diabetic retinopathy-focused program that also screens for referable glaucoma using fundus photography.^17^ We previously reported that among LAC DHS patients who screened positive for referable glaucoma between 2016 to 2018, 25.2% were diagnosed with glaucoma after in-office evaluation.^17^ In the current study, we evaluated the cumulative incidence of treatment initiation and temporal patterns of treatment initiation over an 8-year period after screening and assessed whether baseline characteristics obtained at screening predicted treatment. By examining treatment initiation as an intermediate clinical endpoint indicating clinician-directed efforts to lower IOP, this study provides insight into the long-term clinical impact of glaucoma screening beyond initial diagnostic yield.

## Methods

This study was approved by the Institutional Review Boards of the University of Southern California. The study adhered to the tenets of the Declaration of Helsinki and complied with the Health Insurance Portability and Accountability Act.

### Data Collection

Participants were drawn from the LAC DHS teleretinal screening program, which serves approximately 1,750 incident diabetic patients a month at 17 hospital- and community-based sites.^16^ Participants were screened using dilated fundus photography, performed by trained photographers. During the visit, technicians obtained visual acuity and a brief medical history, including information on diabetic control (self-reported and HbA1c), medications used for diabetes, and the presence of hypertension. Fundus photographs were remotely reviewed by a team of 15 certified optometrists for common eye diseases, including diabetic retinopathy and referable glaucoma. Referable glaucoma was defined at the patient level by a cup-to-disc ratio (CDR) of ≥0.6 or an inter-eye CDR asymmetry ≥0.2. If pathology was identified, referrals were placed for in-office evaluation at LAC DHS eye clinics, where a comprehensive glaucoma evaluation was performed by an ophthalmologist or optometrist.

All patients aged 18 years or older with at least one screening encounter between September 1, 2013 and March 1, 2024 were eligible for this study. In the primary analysis, patients who screened positive for referable glaucoma, received a referral for eye care, and attended at least one in-person visit were included. Participants with record of glaucoma treatment prior to screening were excluded.

Baseline data from the screening encounter were extracted from the screening software (EyePACS, EyePACS LLC, Santa Cruz, California), including screening site, demographics (age, gender, ethnicity, race), glaucoma history (personal or family), medical history (diabetes and hypertension status, need for insulin), baseline visual acuity, and diagnosis of referable glaucoma and diabetic retinopathy by optometrists remotely reviewing the fundus photographs. When visual acuity was available for both eyes, the worse-eye logMAR visual acuity (converted from Snellen) was used to represent baseline visual function at the patient level. Data on eye care visits (dates, completion status), disease diagnoses (ICD-9/10 codes), medication prescriptions (dates, medication names), and glaucoma procedures (CPT codes) were extracted from the LAC DHS electronic health record (Cerner PowerChart, Oracle Health, North Kansas City, Missouri).

Chart-reviewed diagnoses confirmed by glaucoma specialists were available for a subset of the study cohort previously examined in a retrospective chart review, consisting of patients screened between January 1, 2016 and December 31, 2018.^17^ Clinical notes and diagnostic testing (OCT and Humphrey visual field printouts) obtained within one year of the in-office evaluation were reviewed by fellowship-trained glaucoma specialists (B.Y.X, B.J.W.), and patient-level diagnoses of glaucoma, glaucoma suspect, or healthy were assigned.

### Outcomes

The primary outcome of the study was a composite measure of any form of IOP-lowering treatment including topical IOP-lowering medications (based on prescription), selective laser trabeculoplasty (SLT; CPT 65855) or laser peripheral iridotomy (LPI; CPT 66761), and surgical procedures for glaucoma, including trabeculectomy (CPT 66170, 66174, 66179, 66172), glaucoma drainage device or XEN gel stent (CPT 66180, 66183, 65820) implantation, minimally invasive glaucoma surgery (CPT 0449T, 66500, 66625, 0191T, 0671T), and cyclophotocoagulation (CPT 66710, 66711). Surgical treatment was also analyzed individually as a secondary outcome.

Time from screening to treatment initiation was measured. The index date was defined as the date of the first screening encounter resulting in a diagnosis of referable glaucoma and referral for in-office evaluation. Patients were followed until the first treatment event or censored at their last observed eye visit, with follow-up capped at 8 years. Loss to follow-up was defined as a gap between visits exceeding 24 months and censoring was applied at the last visit prior to this interval.

Diagnoses among treated patients were evaluated using two complementary approaches. First, diagnostic codes within a ±12 months window of treatment initiation were aggregated to classify patients as manifest glaucoma (ICD-9 365.1*-365.9*; ICD-10 H40.1-H40.9*, H42.*) or glaucoma suspect (ICD-9 365.0*; ICD-10 H40.0*). Because some coding at LAC DHS is performed by professional coders without clinical knowledge of glaucoma, these codes were used as administrative proxies. To improve specificity, a sub-analysis was performed, restricting to participants with glaucoma specialist-confirmed diagnoses in the aforementioned chart review.^17^

### Statistical Analysis

All analyses were performed at the patient level. Continuous variables were summarized using means and standard deviations, and categorical variables with counts and percentages. Nonparametric cumulative incidence functions (CIFs) were estimated using the Aalen-Johansen method to quantify treatment incidence over time while accounting for censoring and loss to follow-up. Point estimates were reported with 95% confidence intervals.

Cox proportional hazards models were used to evaluate factors associated with glaucoma treatment and loss to follow-up after a positive screening result. Covariates included age, gender, a combined variable representing race and ethnicity (grouped as Asians, Hispanics, Non-Hispanic Blacks, Non-Hispanic Whites, and Other), glaucoma history (personal, family, or none), hypertension, insulin dependence, diabetic retinopathy status (present or absent) at screening, baseline worse-eye visual acuity, and screening site (community or hospital). A time-varying Cox proportional hazards model was used to assess whether treatment initiation was associated with subsequent retention in care, with treatment status included as a time-dependent covariate. Hazard ratios (HRs) with 95% confidence intervals were reported.

Preliminary Cox models suggested that several covariates were associated with both treatment initiation and loss to follow-up, suggesting potential bias if loss to follow-up were treated as non-informative censoring. Loss to follow-up precludes observation of treatment initiation; therefore, it was modeled as a competing risk. Competing risks regression was performed using the Fine-Gray sub-distribution hazards model, with treatment initiation as the event of interest and loss to follow-up as the competing event. The same covariate set was used for modeling. Sub-distribution hazard ratios (SHRs) with 95% CIs were reported, representing the relative effect of covariates on the cumulative incidence of treatment while accounting for the competing risk of loss to follow-up. A secondary analysis was performed with surgical treatment as the outcome. In this analysis, ethnicity categories were collapsed to Hispanic or non-Hispanic and glaucoma history was removed to improve model stability.

Participants were stratified into risk groups based on the linear predictor from the Fine-Gray model and the baseline covariates at screening. For the primary analysis on any treatment, the cohort was divided into tertiles (low, intermediate, and high risk). For the secondary analysis on surgical treatment, the cohort was divided into halves (low and high risk) due to fewer events. CIF curves were generated using the Aalen-Johansen method for each risk group. All statistical analyses were performed using R version 4.4.1 (R Foundation for Statistical Computing, Vienna, Austria). Statistical significance was defined as a two-sided p-value < 0.05.

## Results

The study included 2,367 participants who screened positive for referable glaucoma and attended in-person evaluation from an overall population of 84,716 participants in the screening program. The study cohort had a mean age of 58.5 (SD=10.0) years. Approximately half of the cohort was female (52.8%) and the majority was Hispanic (69.3%). A total of 25.5% had findings of diabetic retinopathy on screening fundus photography. The median duration from screening to in-office evaluation was 21.9 weeks (IQR: 14.0-35.6). Median follow-up was 2.11 years (IQR: 1.21-3.33) (**Table 1**).

**Table 1.** Baseline Demographic and Clinical Characteristics.

| Characteristic |  | Value |
| --- | --- | --- |
| Number of patients |  | 2367 |
| Age & Gender | Age (years) | 58.5 (10.0) |
|  | Female | 1249 (52.8%) |
| Ethnicity | Hispanic | 1641 (69.3%) |
|  | Black | 232 (9.8%) |
|  | Asian | 153 (6.5%) |
|  | White | 39 (1.6%) |
|  | Other | 302 (12.8%) |
| Glaucoma History | None | 2197 (92.8%) |
|  | Family | 112 (4.7%) |
|  | Personal | 58 (2.5%) |
| Comorbidity Status | Hypertension | 1286 (54.3%) |
|  | Insulin Dependent | 546 (23.1%) |
| Screening Findings | Baseline visual acuity (worse eye, logMAR) | 0.3 (0.2) |
|  | Diabetic Retinopathy | 604 (25.5%) |

Results are shown in mean (standard deviation) for continuous variables and n (%) for categorical variables.

During follow-up, 799 participants received treatment and 772 were lost to follow-up. The remaining 796 were monitored without treatment until the study end date. The cumulative incidence of IOP-lowering treatment increased over time from 19.6% (95% CI: 18.0-21.2) in Year 1 to 45.1% (42.1-48.1) by Year 8. The annual incidence was greatest in Year 1 (19.6%), followed by Year 2 (7.5%) and Year 3 (5.5%), after which it remained between 1.2% and 3.4% per year. The cumulative incidence of surgery was substantially lower, ranging from 0.2% (0.0-0.3) in Year 1 to 5.3% (3.4-7.2) in Year 8 with annual incidence remaining low in Years 1 to 4 (0.2-0.7%), then increasing through Years 5 to 8 (0.7-1.5%) (**Figure 1**).

**Figure 1.**
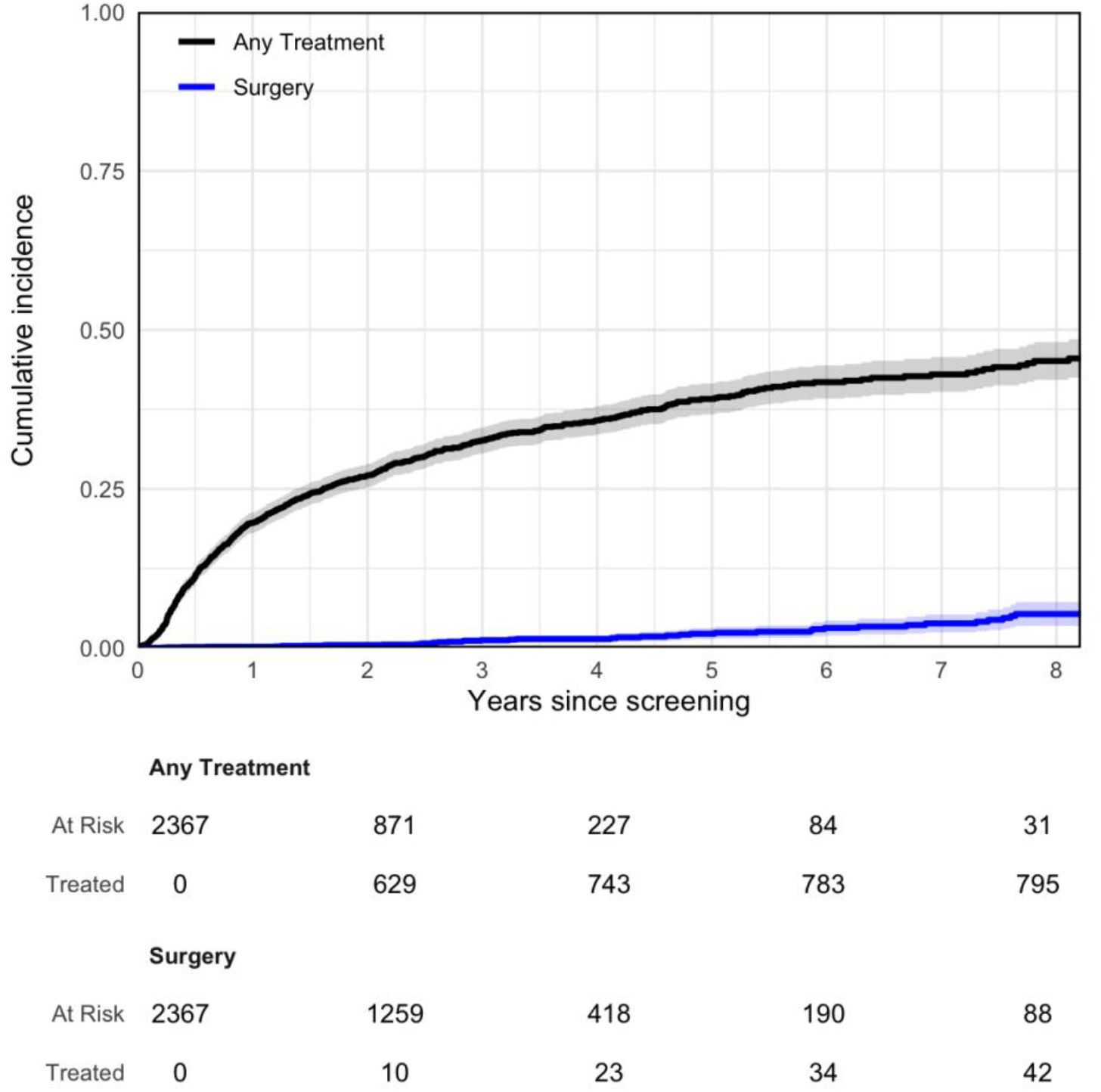
Cumulative Incidence Functions for Treatment and Surgery

In the analysis of the cohort based on diagnostic codes, 2,126 of the 2,367 participants were assigned a diagnostic code within 12 months of the in-office evaluation. Of these, 500 (23.5%) had a glaucoma code, 1,355 (63.7%) had a glaucoma suspect code, and 271 (12.7%) did not have a glaucoma-associated code. The cumulative incidence of IOP-lowering treatment by Year 8 was 80.2% (76.3-84.1) in those with a glaucoma code at baseline and 36.5% (31.6-41.5) in those with a glaucoma suspect code at baseline (**Figure 2**). Treatment incidence in glaucoma cases was concentrated early and plateaued over time, whereas treatment among suspects accumulated progressively throughout follow-up. Similar trends in 8-year cumulative incidence were observed in the sub-analysis of the cohort with chart-reviewed diagnoses (97.1% [93.3-100] in glaucoma; 26.6% [21.5-31.6] in glaucoma suspects).

**Figure 2.**
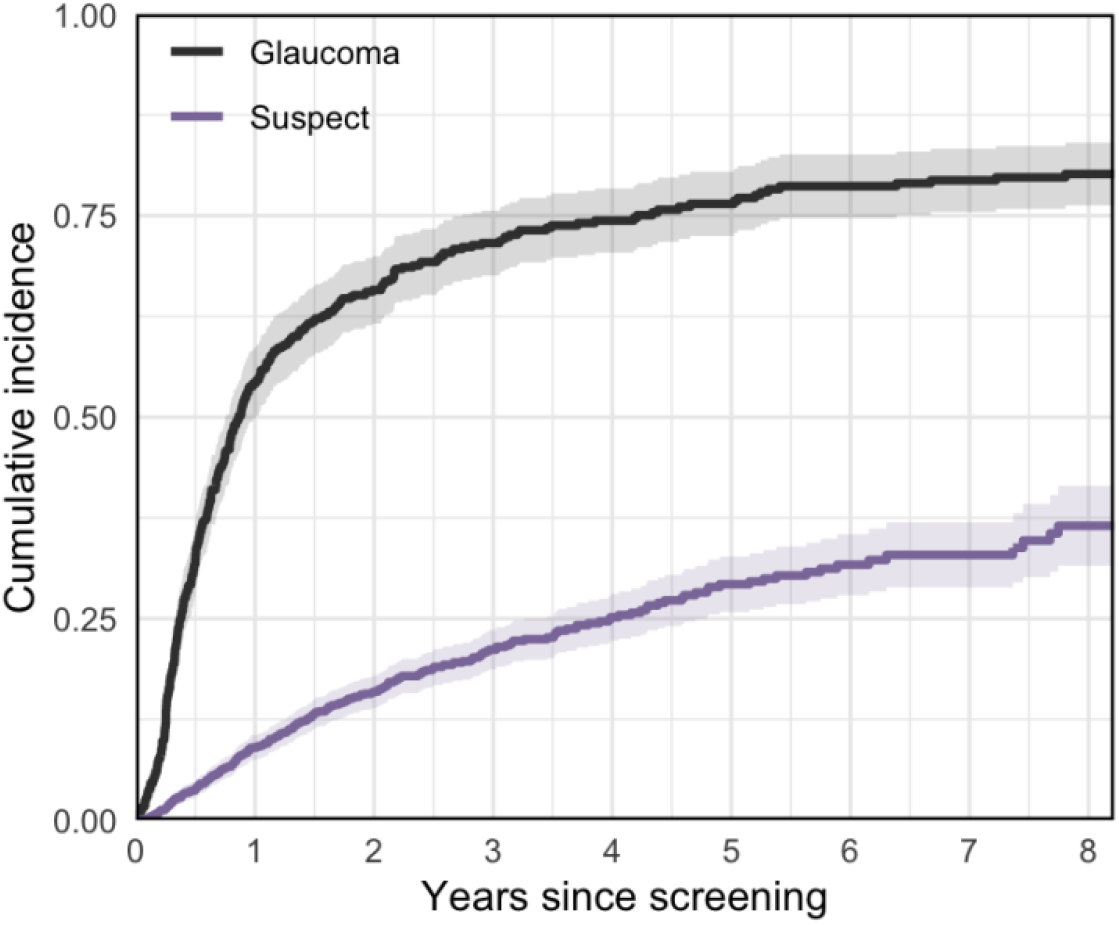
Cumulative Incidence Functions Stratified by Glaucoma Classification

As first-line treatment, 97.5% of participants received medication (n=779), 2.3% received laser (n=18), and 0.2% received surgery (n=2). Prostaglandin analogs were the preferred first-line treatment (87.1%). Among first-line surgeries performed in the study period, MIGS were most common (n=22), followed by implantation of a glaucoma drainage device (n=11) and trabeculectomy (n=9) (**Table 2**).

**Table 2.** Treatment Details.

| Treatment Type |  | n (%) |
| --- | --- | --- |
| First Treatment | Prostaglandin Analog | 696 (87.1%) |
|  | Combination | 50 (6.3%) |
|  | Beta-Blocker | 15 (1.9%) |
|  | Alpha-agonist | 14 (1.8%) |
|  | Peripheral Iridotomy | 14 (1.8%) |
|  | CAI | 4 (0.5%) |
|  | Laser Trabeculoplasty | 4 (0.5%) |
|  | MIGS | 1 (0.1%) |
|  | GDD | 1 (0.1%) |
| First Surgery | MIGS | 22 (51.2%) |
|  | GDD | 11 (25.6%) |
|  | Trabeculectomy | 9 (20.9%) |
|  | CPC | 1 (2.3%) |

Carbonic anhydrase inhibitor (CAI), Minimally invasive glaucoma surgery (MIGS), Glaucoma drainage device (GDD), Cyclophotocoagulation (CPC). Results are shown in n (%) with proportions from within treatment type (any treatment or surgery).

Baseline predictors of treatment were derived from data collected at the screening encounter with Fine-Gray models, handling loss to follow-up as a competing risk to treatment (**Table 3**). Older age (SHR 1.03 [1.02-1.04] per year, p<0.001), Black race (1.50 [1.21-1.87], p<0.001), and personal history of glaucoma (1.90 [1.34-2.70], p<0.001) were risks for initiation of any treatment, and Asian ethnicity was protective (0.71 [0.52-0.97], p=0.03). Family history of glaucoma was a borderline risk (SHR 1.33 [0.98-1.79], p=0.06). Older age (1.06 [1.03-1.09], p<0.001), worse visual acuity (5.11 [2.15-12.12] per logMAR unit, p<0.001), and screening at a hospital-based site (2.46 [1.35-4.49], p=0.003) were associated with greater risk of surgical treatment.

**Table 3.** Fine-Gray Models Predicting All Treatment and Surgery.

| Covariate | Any Treatment |  | Surgery |  |
| --- | --- | --- | --- | --- |
|  | SHR | P Value | SHR | P Value |
| Age (per year) | **1.03 (1.02-1.04)** | **< 0.001** | **1.06 (1.03-1.09)** | **< 0.001** |
| Male (vs. Female) | 1.03 (0.89-1.18) | 0.69 | 1.12 (0.61-2.08) | 0.71 |
| Ethnicity: Asian<br>(vs. Hispanic) | **0.71 (0.52-0.97)** | **0.03** | - | - |
| Ethnicity: Black<br>(vs. Hispanic) | **1.50 (1.21-1.87)** | **< 0.001** | - | - |
| Ethnicity: White<br>(vs. Hispanic) | 1.01 (0.56-1.83) | 0.98 | - | - |
| Ethnicity: Other<br>(vs. Hispanic) | 1.03 (0.83-1.28) | 0.77 | - | - |
| Ethnicity: Non-Hispanic<br>(vs. Hispanic) | - | - | 0.87 (0.45-1.69) | 0.68 |
| Glaucoma History: Family<br>(vs. None) | 1.33 (0.98-1.79) | 0.06 | - | - |
| Glaucoma History: Personal<br>(vs. None) | **1.90 (1.34-2.70)** | **< 0.001** | - | - |
| Hypertension | 1.07 (0.93-1.24) | 0.36 | 0.79 (0.43-1.45) | 0.44 |
| Insulin Dependent | 0.97 (0.82-1.15) | 0.74 | 0.93 (0.44-1.99) | 0.86 |
| Diabetic Retinopathy | 0.99 (0.84-1.16) | 0.87 | 0.84 (0.41-1.73) | 0.64 |
| Baseline visual acuity (worse eye,<br>logMAR) | 1.27 (0.93-1.73) | 0.13 | **5.11 (2.15-12.12)** | **< 0.001** |
| Screening Site: Hospital-Based<br>Center (vs. Community) | 0.88 (0.76-1.02) | 0.09 | **2.46 (1.35-4.49)** | **0.003** |

Fine-Gray subdistribution hazards ratios (SHR) for the initiation of all treatment and surgical treatment are shown with 95% confidence intervals. Loss to follow-up (LTFU) was handled as a competing risk. Results with significant *p*-values are shown bold.

Risk stratification based on model-derived predictors demonstrated substantial separation in cumulative incidence curves (**Figure 3**). After 8 years of follow-up, the cumulative incidence of treatment was 59.9% (54.7-65.2%) in the high-risk tertile versus 31.2% (26.3-36.2%) in the low-risk tertile, and for surgery, was 9.7% (6.1-13.3%) in the high-risk half versus 1.0% (0.0-2.0%) in the low-risk half.

**Figure 3.**
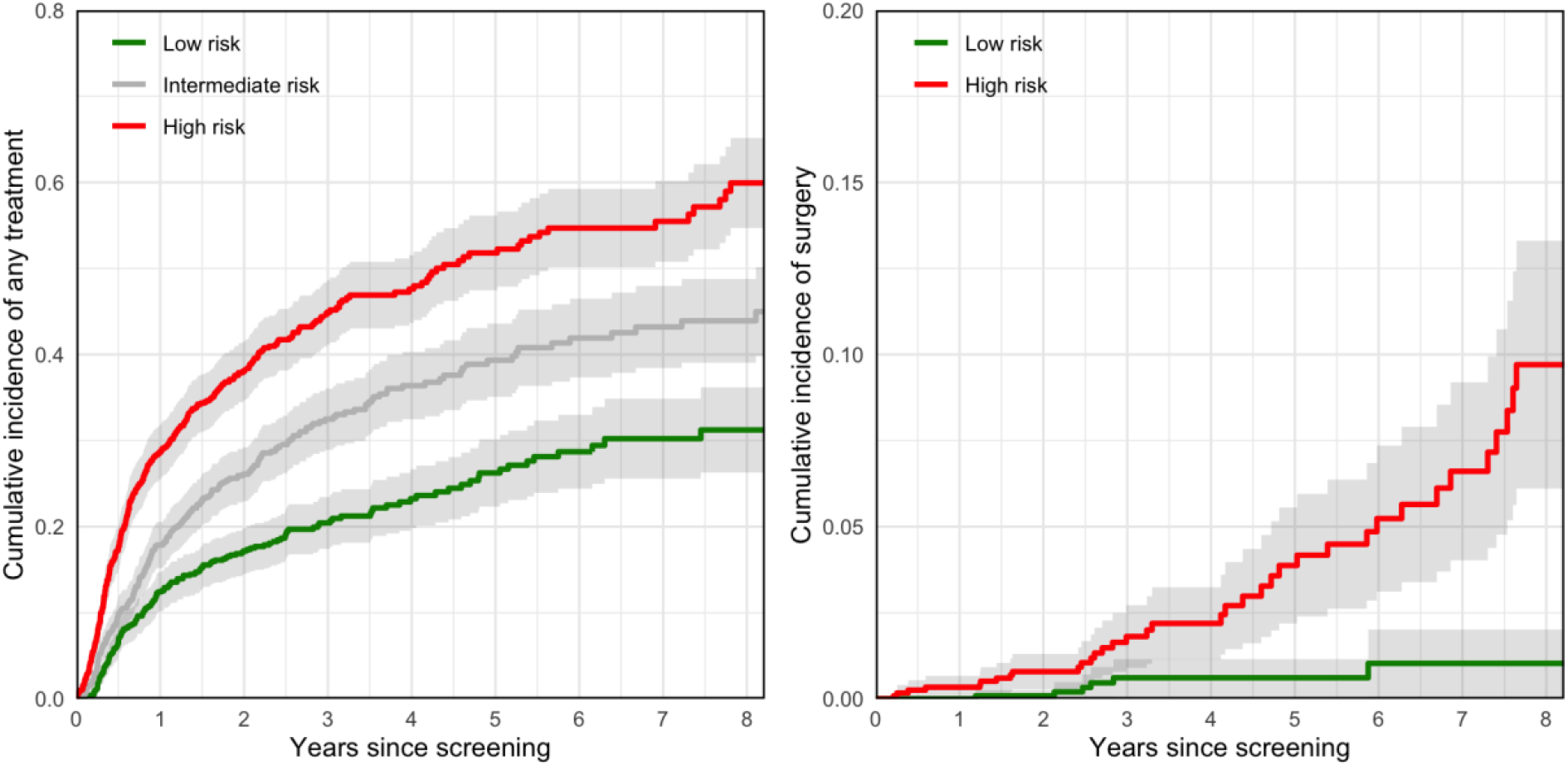
Cumulative Incidence Functions Stratified by Risk for Any Treatment and Surgery

In analysis of factors associated with loss to follow-up, Black race (HR 1.29 [1.06-1.57], p=0.01), insulin dependence (HR 1.16 [1.00-1.34], p=0.05), and worse baseline visual acuity (HR 1.31 [1.01-1.70], p=0.04) were associated with greater risk. In the time-varying Cox proportional hazards model, receiving IOP-lowering treatment was associated with a 20.1% reduction in the hazard of subsequent loss to follow-up (HR 0.80 [0.68-0.94], p=0.008) (**Supplemental Table 1**).

## Discussion

IOP-lowering treatment was initiated in nearly half of safety-net diabetic patients who screened positive for referable glaucoma over an 8-year period. Treatment initiation was concentrated in the early period in patients initially diagnosed as glaucoma, but rates steadily increased over time, reflecting treatment initiation among patients initially diagnosed as glaucoma suspects. Surgical intervention was comparatively uncommon but increased steadily over time. Baseline characteristics available at screening were associated with treatment and enabled risk stratification of long-term treatment burden. These findings highlight that a substantial proportion of safety-net diabetic patients who screen positive for referable glaucoma ultimately receive intervention and that early clinical features can identify higher-risk subgroups. By extending beyond cross-sectional diagnostic endpoints, this study addresses an evidence gap by characterizing longitudinal clinical outcomes following screening in a high-risk population, using treatment initiation as an intermediate endpoint for clinician-assessed risk and intent to lower IOP—an outcome aligned with USPSTF priorities.^13^

There is currently a lack of longitudinal evidence regarding the effects of glaucoma screening, as underscored by the USPSTF.^13^ We selected IOP-lowering treatment as a pragmatic longitudinal endpoint that reflects clinical decision-making. It serves as an intermediate endpoint aligned with USPSTF-identified outcomes, which include IOP, structural and functional damage, and patient-reported outcomes.^13^ While initiation of IOP-lowering treatment does not directly measure IOP reduction or downstream structural or functional outcomes, it represents the clinical decision to intervene to achieve these goals. It is also readily observable in longitudinal real-world datasets and is less susceptible to miscoding. Glaucoma status, evaluated using both administrative codes and chart-reviewed diagnoses, supports the validity of this endpoint, as treatment incidence was highest and occurred earliest among patients diagnosed with glaucoma, although treatment was also initiated in glaucoma suspects over time. Therefore, IOP-lowering treatment represents a pragmatic and clinically grounded endpoint for evaluating the longitudinal impact of screening in real-world populations.

Our primary analysis intentionally focused on screen-positive patients who completed in-office evaluation, allowing isolation of outcomes attributable to glaucoma-specific screening rather than broader program-level factors. Within this cohort, the cumulative incidence of treatment was 19.6% in the first year, consistent with prior screening studies, including the SToP Glaucoma Program, Philadelphia Telemedicine Glaucoma Detection and Follow-up Study, and Manhattan Vision Screening Study, which diagnosed manifest glaucoma in 11-34% of evaluated participants. Treatment was initiated in approximately 11-50% of participants within the first year after evaluation, although direct comparisons should be interpreted cautiously given differences in referral criteria, disease and outcome definitions, and treatment thresholds.^15,18,19^ Nevertheless, treatment initiation reflects both disease severity and provider decision-making, providing a meaningful measure of the clinical impact of screening. These variations underscore the need for standardized outcome definitions and reporting frameworks in glaucoma screening research.

Surgical intervention was uncommon in this cohort, with a cumulative incidence of 5.3% at 8 years. The longitudinal incidence of surgery after population-based glaucoma screening has not been well characterized. Our observed cumulative incidence is lower than the 19.5% cumulative incidence of procedural intervention over 4 years reported in patients with primary open angle glaucoma in the IRIS registry, although 63.5% of the events were laser trabeculoplasty, and differences in patient populations and outcome definitions limit direct comparison.^20^ Overall, the low incidence of surgical intervention suggests that screening may identify earlier-stage disease, although surgical decision-making is also affected by practice patterns, disease severity, and access to subspecialty care.

Our extended longitudinal follow-up revealed a temporal pattern in treatment initiation not well characterized in prior screening studies. Treatment initiation was most frequent in the first year after screening but continued to accumulate over time with progressively smaller annual increases. Stratification by glaucoma classification after in-office evaluation demonstrated that treatment in glaucoma cases occurred early and plateaued over time, whereas treatment among glaucoma suspects accumulated gradually throughout follow-up, reaching approximately 36.5% by Year 8. This finding suggests that screen-positive patients initially classified as glaucoma suspects contribute meaningfully to long-term treatment burden and warrant ongoing surveillance. At the same time, most glaucoma suspects remained untreated, highlighting the need for improved risk stratification approaches to identify those at greatest risk of future treatment. Notably, the sustained annual incidence of treatment after the initial period is consistent with reported conversion rates from glaucoma suspects to glaucoma and with incident rates observed in the Ocular Hypertension Treatment Study (OHTS).^21,22^ These findings indicate that the impact of screening extends beyond initial diagnosis and should be evaluated in terms of both detection of prevalent disease and identification of individuals at risk for future progression.

Baseline factors assessed at screening demonstrated meaningful associations with treatment and enabled risk stratification of long-term treatment burden. The Fine-Gray model separated the cumulative incidence of treatment between risk groups, with a 28.7% difference at 8 years, most of which accrued in the first year after screening. This degree of separation is comparable to the OHTS risk model, which has been proposed for guiding the timing of examinations and preventive treatment in ocular hypertensives.^22^ In this context, high-risk individuals could be prioritized for expedited evaluation or educated about the importance of follow-up, whereas lower-risk individuals could be managed with targeted re-screening strategies. The current model, however, should be considered proof of concept, as the cumulative incidence remains non-trivial in the low-risk group. Our preliminary model for treatment relied on established glaucoma risk factors, including age, race, and personal and family history, while the model for glaucoma surgery demonstrated additional effects consistent with disease severity, cataract status, and access to specialty care.^1,23,24^ Importantly, several of these effects—Black race and worse visual acuity—were associated with both treatment initiation and loss to follow-up, indicating that patients at high risk for needing treatment may also be at high risk for loss to follow-up. While treatment was associated with retention, loss to follow-up should not be interpreted as the absence of treatment need. High attrition rates emphasize the need for approaches that combine risk stratification with targeted interventions to improve retention in care.^25–27^

This study has some limitations. First, our results may not be generalizable to the broader U.S. population due to the demographic composition, primary focus on diabetic retinopathy by the LAC DHS screening program, and the high screening yield of glaucoma; however, they provide robust, longitudinal evidence on a large safety-net screening cohort, with analyses designed to isolate glaucoma-specific effects and provide generalizable insights in a high-risk screening population. Second, attrition was substantial and, despite being modeled as a competing risk, may have affected the accuracy of cumulative incidence estimates. Further work is needed to understand the determinants of loss to follow-up and identify effective interventions to mitigate it. Third, our study cohort was selected from participants completing in-clinic evaluation, which may have introduced selection bias for participants more likely to engage in care or receive treatment. Fourth, risk stratification was limited by the absence of imaging-derived features from baseline fundus photography (e.g., cup-to-disc ratio, thinning of the neuroretinal rim), which may improve predictive performance and should be incorporated in future work using expert grading or deep learning approaches.^28^ Finally, reliance on diagnostic coding to infer treatment for glaucoma introduces potential misclassification, although this was partially mitigated through a chart-reviewed sub-analysis confirming that treatment was primarily initiated for glaucoma. Future work will focus on developing more robust glaucoma phenotypes using structured health record data and diagnostic testing.

In conclusion, this study provides longitudinal characterization of IOP-lowering treatment in a large cohort screening positive for referable glaucoma in a teleretinal screening program. Treatment in this high-risk population was common, increased over time, and varied meaningfully by baseline risk assessment. These findings suggest that glaucoma screening programs influence care beyond the initial diagnostic evaluation and that early clinical features may help prioritize patients for timely follow-up and management. Future work integrating imaging-derived features, diagnostic data, and social determinants of health may help identify glaucoma suspects at greatest risk for future treatment and optimize care pathways following screening. Additional studies evaluating the long-term effects of treatment initiation on IOP and structural and functional outcomes may further inform evidence-based policy regarding glaucoma screening.

## Data Availability

All data produced in the present work are contained in the manuscript

## Acknowledgements

This work was supported by the National Center for Advancing Translational Science (NCATS) of the National Institutes of Health under award number KL2TR001854 and UL1TR001855, and R01 EY035677 and K23 EY032985 from the National Eye Institute, National Institutes of Health, Bethesda, Maryland. The content is solely the responsibility of the authors and does not necessarily represent the official views of the National Institutes of Health. The work was additionally supported by a Mentoring for Advancement of Physician-Scientists (MAPS) Award from the American Glaucoma Society; a DHS-USC Safety Net Innovation Award from the Southern California Clinical and Translational Science Institute; a AI4Health Award from the University of Southern California; and an unrestricted grant to the Department of Ophthalmology from Research to Prevent Blindness, New York, NY.

## Supplement

**Supplemental Table 1.** Cox Proportional Hazards Model for Loss to Follow-Up.

| **Covariate** | **Hazard Ratio** | ***P***<br>**Value** |
| --- | --- | --- |
| Age (per year) | 0.99 (0.99-1.00) | 0.13 |
| Male (vs. Female) | 1.09 (0.96-1.23) | 0.17 |
| Ethnicity: Asian (vs. Hispanic) | 1.15 (0.90-1.46) | 0.26 |
| Ethnicity: Black (vs. Hispanic) | **1.29 (1.06-1.57)** | **0.01** |
| Ethnicity: White (vs. Hispanic) | 0.79 (0.47-1.32) | 0.36 |
| Ethnicity: Other (vs. Hispanic) | 0.91 (0.75-1.11) | 0.34 |
| Glaucoma History: Family (vs. None) | 0.99 (0.75-1.31) | 0.96 |
| Glaucoma History: Personal (vs. None) | 0.94 (0.64-1.38) | 0.76 |
| Hypertension | 1.01 (0.89-1.14) | 0.92 |
| Insulin Dependent | **1.16 (1.00-1.34)** | **0.05** |
| Diabetic Retinopathy | 0.87 (0.75-1.01) | 0.07 |
| Baseline visual acuity (worse eye, logMAR) | **1.31 (1.01-1.70)** | **0.04** |
| Screening Site: Academic Center (vs. Community) | 1.06 (0.94-1.21) | 0.33 |
| Treatment | **0.80 (0.68-0.94)** | **0.008** |

Hazards ratios for becoming lost to follow-up are shown with 95% confidence intervals. Treatment was handled as a time-varying effect. Results with significant *p*-values are shown bold.

